# Adherence to a healthy lifestyle and brain structural imaging markers

**DOI:** 10.1101/2022.08.12.22278716

**Authors:** Yuesong Pan, Jie Shen, Xueli Cai, Hui Chen, Geng Zong, Wanlin Zhu, Jing Jing, Tao Liu, Aoming Jin, Yilong Wang, Xia Meng, Changzheng Yuan, Yongjun Wang

## Abstract

**Objective:** To investigate the association of adherence to a healthy lifestyle with a panel of brain structural markers in middle-aged and older adults.

**Design:** Cross-sectional and prospective study design.

**Setting:** PolyvasculaR Evaluation for Cognitive Impairment and vaScular Events (PRECISE) study in China and UK Biobank (UKB).

**Participants:** 2,413 participants in PRECISE and 19,822 participants in UKB.

**Exposures:** A healthy lifestyle score (0-5) was constructed based on five modifiable lifestyle factors: healthy diet, physically active, non-current-smoking, non-alcohol consumption (in PRECISE)/moderate alcohol consumption (in UKB), and healthy body weight.

**Main Outcomes:** Validated multimodal neuroimaging markers were derived from brain magnetic resonance imaging (MRI).

**Results:** In the cross-sectional analysis of PRECISE, participants who adopted four or five low-risk lifestyle factors had larger total brain volume (TBV; β= 0.12, 95%CI: - 0.02, 0.26; *p-trend* = 0.048) and gray matter volume (GMV; β= 0.16, 95%CI: 0.01, 0.30; *p-trend* = 0.047), smaller white matter hyperintensity volume (WMHV; β= -0.35, 95%CI: -0.50, -0.20; *p-trend* <0.001) and lower odds of lacune (Odds Ratio [OR]=0.48, 95%CI: 0.22, 1.08; *p-trend* = 0.03), compared to those with zero or one low-risk factors. Meanwhile, in the prospective analysis in UKB (with a median of 7.7 years’ follow-up), similar associations were observed between the number of low-risk lifestyle factors (4-5 vs 0-1) and TBV (β= 0.22, 95%CI: 0.16, 0.28; *p-trend* <0.001), GMV (β= 0.26, 95%CI: 0.21, 0.32; *p-trend* < 0.001), white matter volume (WMV; β= 0.08, 95%CI: 0.01, 0.15; *p-trend* = 0.001), hippocampus volume (β= 0.15, 95%CI: 0.08, 0.22; *p-trend* = <0.001), and WMHV burden (β= -0.23, 95%CI: -0.29, -0.17; *p-trend* < 0.001). Those with four or five low-risk lifestyle factors showed approximately 2.0-5.8 years of delay in aging of brain structure.

**Conclusion:** Adherence to a healthier lifestyle was associated with a lower degree of neurodegeneration-related brain structural markers in middle-aged and older adults.

**What is already known on this topic:** Previous research has linked specific modifiable lifestyle factors to age-related cognitive decline in adults.

Little is known about the potential role of an overall healthy lifestyle in brain structure.

**What this study adds:** In the cross-sectional analysis of 2,413 participants in China and the prospective analysis of 19,822 participants in UK, participants who adopted 4-5 low-risk lifestyle factors had larger total brain volume and gray matter volume and lower white matter hyperintensity volume, compared to those with 0-1 factors.

The association estimates were equivalent to approximately 2.0-5.8 years of delay in aging of brain structure.

Adherence to a healthier lifestyle was associated with a lower degree of neurodegeneration-related brain structural markers in middle-aged and older adults.

## Introduction

The continuous increment in life expectancy is accompanied by rising prevalence of brain ageing and neurological disability, such as dementia^1 2^, and the prevalence is projected to dramatically increase over the next three decades^3^. As sensitive precursors of preclinical stage of dementia, neuroimaging brain structural markers have been increasingly utilized to investigate risk factors of the aging brain and related underlying mechanisms. Given the public priority to the formulation of effective preventive strategies, it is, therefore, essential to understand the risk factors for neurodegeneration-related brain structural markers.

Increased attention has been focused on a constellation of novel lifestyle factors, including diet quality, cigarette smoking, physical activity, alcohol consumption, and body weight^4 5^. Recent studies have proposed that long-term lifestyles may induce alterations within the brain in older adult life, which potentially act via atherosclerotic processes, neurotrophic factors, and chronic diseases consequences^6^. In observational studies, several healthy lifestyle factors have been linked to a lower brain atrophy separately^7-18^. However, in real life, many of these factors are interrelated, yet few studies have examined the lifestyle factors in combination with brain structural markers^19 20^. The generalizability of the findings from existing studies might be limited by small sample size, suboptimal control for important confounders, or both. Thus, large-scale studies are warranted to elucidate whether different behaviors cumulatively and simultaneously influence late-life brain health.

We therefore used the large sample of neuroimaging data and detailed assessments of lifestyle factors from two independent population-based studies, the PolyvasculaR Evaluation for Cognitive Impairment and vaScular Events (PRECISE) study in China and UK Biobank (UKB) in the UK, to examine the association of adherence to a healthy lifestyle with a panel of neurodegeneration brain structural markers.

## Methods

### Study Population

The PRECISE study is an ongoing population-based cohort of 3,067 dementia-free adults aged 50–75 years sampled from six villages and four communities of Lishui city, southeast of China. Participants were enrolled and then performed comprehensive face-to-face interviews, physical evaluation, and brain MRI, between May 2017 and September 2019. Further details of the study have been described elsewhere^21^. The UK Biobank (UKB) is a prospective cohort study of over 500,000 community-dwelling participants aged 40-69 years across the United Kingdom, since 2006-2010^22^. Extensive information was collected at recruitment and the brain MRI scan was performed since 2014.

In PRECISE, we excluded participants with a history of stroke (n=87) and missing information on brain MRI measures (n=567). The final cross-sectional analyses included 2,413 participants. In UKB, among the 20,200 participants who underwent structural MRI brain scan, we excluded 162 individuals who had prevalent dementia or stroke via hospital inpatient records and 216 individuals who had missing data on BMI, alcohol consumption, smoking status, physical activity, and diet. The final analytical set included 19,822 participants (Supplementary Fig 1).

### Assessment of Lifestyle Factors

Based on research evidence and expert knowledge on the health benefits of lifestyle factors in brain health^23^, we selected five modifiable lifestyle factors - diet, physical activity, smoking status, alcohol consumption, and body mass index (BMI).

Information on these lifestyle factors was collected from self-reported questionnaires or physical examination and then dichotomized according to prespecified cutoffs (Supplementary Table 1). Although diverse populations were enrolled and inconsistent assessment methods were used in PRECISE and UKB, we applied study-specific definitions of certain lifestyle factors appropriate for Western and Asian populations, respectively. A healthy lifestyle score was computed by summing low-risk lifestyle factors on a scale of 0-5, with higher scores indicating better adherence to healthier lifestyle.

In the PRECISE, information on dietary intake was assessed using a simplified self-reported food frequency questionnaire (FFQ), related to the six major food groups: red meat, poultry, aquatic products, eggs, fresh vegetables, and fresh fruits. Participants were asked how often and amount, they consumed specific foods on a normal day. The reliability and validity of the dietary quality generated using simplified FFQ for Chinese have been described previously^24^. Dietary quality was assessed using the dietary diversity score (DDS). We scored 1 point for an individual who consumed any food group no less than once per day, 0 points otherwise; and a total of 6 points of DDS represented the highest level of dietary diversity^25 26^. A healthy diet was defined as the higher DDS in the top 20% of cohort distribution (scores 4-6). For physical activity, participants were asked the time spent in vigorous activities or moderate activities during a usual week and we then calculated the daily metabolic equivalent hours of physical activity^27^. Physically active was defined as the metabolic equivalent in the upper quartile. Participants were categorized as current and non-current smokers, and the later was considered as the low-risk group. Previous study pointed out that potential protective role of alcohol drinking on cognitive performance may have a relationship with wine type^28^. Given that older adults in China prefer white wine with a higher alcohol content, we defined non-alcohol consumption as the low-risk group in PRECISE. Body weight and height were measured by trained medical staff. We defined as the body mass index (BMI; weight in kilograms divided by height in meters squared) in the range of 18.5 - 24.0 kg/m^2^ specific for Chinese^29^.

In the UKB, dietary data were obtained using a simplified FFQ and dietary quality was evaluated according to the criteria as adequate consumption of 4 healthy food groups (fruits, vegetables, fish, whole grains) and reduced consumption of 3 food groups (refined grains, processed meats, and unprocessed red meats), following dietary recommendations of the American heart Association. Healthy diet was defined as meeting at least 4 items of the dietary recommendations^23^. Physically activity was defined as at least 150 minutes of moderate activity per week or 75 minutes of vigorous activity per week (or an equivalent combination) or engaging in moderate physical activity at least 5 days a week or vigorous activity once a week, which was following the American Heart Association recommendations^23^. Consistent with the definition of PRECISE study, non-current smoking was conceived as a low-risk lifestyle. For alcohol consumption, according to the previous studies in the UK Biobank, moderate alcohol consumption (>0-14 g/d for women and >0-28 g/d for men) was defined as a low-risk level^23^. Healthy body weight was defined as the BMI in the range of 20.0 - <25.0 kg/m^2^, following the World Health Organization (WHO) classification^30^.

### Measurement of Neuroimaging Markers

All brain structural markers utilized in this study were obtained from magnetic resonance imaging (MRI). The neuroimaging markers included brain structural markers (such as total brain volume [TBV], gray matter volume [GMV], white matter volume [WMV], hippocampus volume, white matter hyperintensities volume [WMHV], and lacune). The TBV was calculated as the sum of GWM and WMV.

All participants were scanned on the same MRI scanner at the Lishui Hospital Medical Centre in China and Cheadle Manchester Centre in UK. Briefly, structural MRI data were processed applying a pipeline to the T1 images that used gradient distortion correction, field of view reduction, registration to the standard atlas, brain extraction, defacing, and finally segmentation. In PRECISE, each T1 weighted images was processed using FreeSurfer default processing pipeline (version 7.0) and WMHV data was summarized applying White matter Hyperintensities Analysis Tools (WHAT) software^31^; meanwhile, corresponding imaging variables in UKB study were derived from the image-derived phenotypes (IDPs) released by the UK Biobank team^32^. In PRECISE, lacune of presumed vascular origin, as a marker of cerebral small vessel disease, was defined as rounded or ovoid lesion in the subcortical, BG, or brain stem, with diameter ranging from 3 to 15 mm and cerebrospinal fluid signal density on T2 and FLAIR sequences and no increased signal on DWI^33^.

To correct for differences in head size across participants, we used the residual method (regression-based predicted brain tissue volumes run with intracranial volume [ICV], as a proxy for head size) in PRECISE^34^; correspondingly, head-size normalized volumes of brain regions were also released by UKB team. All brain structural markers were standardized using z-transformation based on the mean and SD for each region separately. WMHV was log-transformed before being z-standardized because of its right-skewed distribution.

### Covariates

Detailed information on sociodemographic characteristics was collected through self-reported questionnaires, including age, sex, ethnicity, type of residence, marital status, and educational level. Information with respect to a medical history of comorbidities (including hypertension, diabetes mellitus, heart disease, tumor/cancer, or dyslipidemia) was collected through either self-reported diagnosis history or determined through medical examinations, hospital medical records, and cancer registry. In PRESICE, we additionally performed cognitive screening test modeled on the Montreal Cognitive Assessment (MoCA), a validated clinical algorithm for risk of cognitive decline^35^. Scores ranged from 0 to 30 points, with a higher score indicating higher cognitive function.

### Statistical Analysis

We categorized the study population as 0-1, 2-3, and 4-5 low-risk lifestyle factors. Characteristics of participants by the number of low-risk lifestyle factors were compared using the one-way analysis of variance (ANOVA) or Kruskal-Wallis test for continuous variables and χ^2^ test for categorical variables.

In the primary analysis, we used general linear models and logistic regression models to examine the association of the number of healthy lifestyle factors with brain structural markers, including TBV, GMV, WMV, hippocampal volume, WMHV, and categorical lacune (only in PRECISE). Multivariable models were adjusted for age, square of age, sex, ethnicity, type of residence, marital status, and educational levels. The *P* values for linear trend were computed by modeling healthy lifestyle score as a continuous variable. In the secondary analysis, we examined which individual lifestyle factor drove the relationship between the number of low-risk lifestyle factors and brain structural markers with additionally mutual adjustment for the other lifestyle factors. We performed several stratified analyses and sensitivity analyses to test the robustness of the results.

We performed several sensitivity analyses to test the robustness of the results. Since health conditions may lie within the causal pathway between lifestyle behaviors and brain structural markers, we further adjusted for the history of major comorbidities (i.e., hypertension, heart disease, diabetes mellitus, tumors, or dyslipidemia). We also excluded participants who had a history of above comorbidities, leaving relatively healthier populations at enrollment. In addition, to control the influence of definition of healthy diet in PRECISE, we repeated the primary analysis using a modified healthy lifestyle score in which redefining the healthy diet as a diet rich in vegetables and fruits (consumed everyday) and limited in red meat (consumed 1 to 6 days a week)^29^. To address the concern about the controversial roles of alcohol consumption associated with brain health, we conducted separate analysis using another modified healthy lifestyle score that was based on the other 4 healthy factors without regard to alcohol. Meanwhile, in UKB, we also redefined the healthy body weight as BMI in the range of 18.5 - <25.0 kg/m^2^, consistent with previous studies in UKB; and redefined the low-risk level of alcohol consumption as non-current alcohol consumption. Lastly, we assessed the association of the number of low-risk lifestyle factors with MoCA score using GLMs in PRECISE.

Data were analyzed with the use of SAS software, Version 9.4 (in PRECISE analysis) and R 3.6.3 (in UKB analysis), with a two-sided P value less than 0.05 indicating statistical significance.

## Results

### Characteristics of the study population

Among 2431 dementia-free participants in PRECISE (mean age 61.3±6.6 years, 53.9% female), the number of participants who adopted 0-1, 2-3, and 4-5 low-risk lifestyle factors were 302 (12.5%), 1735 (71.9%), and 376 (15.6%), respectively (Table 1). Participants with zero or one low-risk lifestyle factor were more likely to be male, illiterate, have higher prevalence of hypertension and diabetes mellitus. In UKB, a total of 19822 participants (mean age 54.83±7.47 years, 47.4% female) at baseline were included with MRI assessment up to 13 years later (median [IQR] = 7.7 [6.7-8.8] years) (Table 2). Among them, 804 (4.1%) had 0-1 low-risk lifestyle factors, 9418 (47.5%) had 2-3 lifestyle factors, and 9600 (48.4%) had 4-5 lifestyle factors. Those with low adherence to a healthy lifestyle were more likely to be female, live in urban areas, have lower education level and higher prevalence of hypertension, diabetes mellitus, and dyslipidemia.

**Table 1.** Characteristics of participants in PRECISE.

| Variables | Total | No. of low-risk lifestyle factors |  |  |  |
| --- | --- | --- | --- | --- | --- |
|  |  | Zero or one | Two or three | Four or five | P-value |
| No. | 2413 | 302 | 1735 | 376 |  |
| Age, y, mean [SD] | 61.3±6.6 | 61.3±6.3 | 61.6±6.6 | 60.0±6.3 | <0.001 |
| Sex, female, n (%) | 1300 (53.9) | 20 (6.6) | 1007 (58.0) | 273 (72.6) | <0.001 |
| Han ethnicity, n (%) | 2329 (96.5) | 287 (95.0) | 1682 (96.9) | 360 (95.7) | 0.17 |
| Type of residence, n (%) |  |  |  |  | 0.22 |
| Urban | 1478 (61.3) | 173 (57.3) | 1065 (61.4) | 240 (63.8) |  |
| Rural | 935 (38.7) | 129 (42.7) | 670 (38.6) | 136 (36.2) |  |
| Marital status, n (%) |  |  |  |  | 0.02 |
| Married | 2199 (91.1) | 284 (94.0) | 1564 (90.1) | 351 (93.4) |  |
| Not married, separated, divorced, and others | 214 (8.9) | 18 (6.0) | 171 (9.9) | 25 (6.6) |  |
| Education, n (%) |  |  |  |  | 0.003 |
| Illiteracy | 387 (16.0) | 30 (9.9) | 306 (17.6) | 51 (13.6) |  |
| Primary school | 594 (24.6) | 90 (29.8) | 429 (24.7) | 75 (19.9) |  |
| Junior school | 733 (30.4) | 93 (30.8) | 514 (29.6) | 126 (33.5) |  |
| High school | 511 (21.2) | 67 (22.2) | 350 (20.2) | 94 (25.0) |  |
| College school | 188 (7.8) | 22 (7.3) | 136 (7.8) | 30 (8.0) |  |
| Hypertension, n (%) | 1048 (43.4) | 156 (51.7) | 771 (44.4) | 121 (32.2) | <0.001 |
| Diabetes mellitus, n (%) | 537 (22.3) | 61 (20.2) | 419 (24.1) | 57 (15.2) | <0.001 |
| Heart disease, n (%) | 211 (8.7) | 26 (8.6) | 152 (8.8) | 33 (8.8) | >0.99 |
| Tumor, n (%) | 315 (13.1) | 12 (4.0) | 241 (13.9) | 62 (16.5) | <0.001 |
| Dyslipidemia, n (%) | 524 (21.7) | 59 (19.5) | 399 (23.0) | 66 (17.6) | 0.04 |
| Healthy diet (DDS = [4, 5, 6]), n (%) | 471 (19.5) | 6 (2.0) | 244 (14.1) | 221 (58.8) | <0.001 |
| Physically active (Q4), n (%) | 592 (24.9) | 25 (8.6) | 334 (19.5) | 233 (62.5) | <0.001 |
| Non-current-smoking, n (%) | 1932 (80.1) | 78 (25.9) | 1483 (85.4) | 371 (98.6) | <0.001 |
| Non-alcohol consumption, n (%) | 1976 (81.9) | 76 (25.2) | 1529 (88.2) | 371 (98.6) | <0.001 |
| Healthy body weight, (BMI = 18.5 - 24.0 kg/m<sup>2</sup>), n (%) | 1229(50.9) | 73(24.2) | 811(46.7) | 345(91.8) | <0.001 |
| Brain structural markers |  |  |  |  |  |
| TBV<sup>a</sup>, ml, mean [SD] | 1032.4±41.4 | 1027.6±45.1 | 1031.7±41.6 | 1039.4±36.5 | 0.002 |
| GMV<sup>a</sup>, ml, mean [SD] | 580.0±25.0 | 576.5±26.6 | 580.0±25.0 | 582.7±23.3 | 0.01 |
| WMV<sup>a</sup>, ml, mean [SD] | 452.4±28.2 | 451.1±32.6 | 451.7±28.0 | 456.7±24.6 | 0.007 |
| Hippocampus<sup>a</sup>, ml, mean [SD] | 8.07±0.66 | 8.05±0.69 | 8.06±0.65 | 8.13±0.65 | 0.21 |
| WMHV<sup>a</sup>, ml, median [IQR] | 1.51 (0.60-3.48) | 1.90 (0.80-4.58) | 1.54 (0.61-3.47) | 1.13 (0.42-2.77) | <0.001 |
| Lacune, n (%) | 107 (4.4) | 22 (7.3) | 75 (4.3) | 10 (2.7) | 0.01 |

<sup>a</sup> Brain tissue volumes were normalized using the residual method to correct the intracranial volume (ICV), as a proxy for head size.
BMI denotes body mass index; DDS denotes dietary diversity score; Q denotes quartile; TBV denotes total brain volume; GMV denotes gray matter volume; WMV denotes white matter volume; WMHV denotes white matter hyperintensity volume.

**Table 2.** Baseline characteristics of participants in UKB.

| Variables | Total | No. of low-risk lifestyle factors |  |  | P-value |
| --- | --- | --- | --- | --- | --- |
|  |  | Zero or one | Two or three | Four or five |  |
| No. | 19822 | 804 | 9418 | 9600 |  |
| Age, y, mean [SD] | 54.83 (7.5) | 53.81 (7.2) | 54.74 (7.4) | 55.02 (7.5) | <0.001 |
| Sex, female, n (%) | 9396 (47.4) | 484 (60.2) | 4935 (52.4) | 3977 (41.4) | <0.001 |
| White Ethnicity, n (%) | 19248 (97.1) | 776 (96.5) | 9149 (97.1) | 9323 (97.1) | 0.59 |
| Type of residence, n (%) |  |  |  |  | <0.001 |
| Urban | 16581 (83.6) | 697 (86.7) | 7976 (84.7) | 7908 (82.4) |  |
| Rural | 3241 (16.4) | 107 (13.3) | 1442 (15.3) | 1692 (17.6) |  |
| Marital status, n (%) |  |  |  |  | 0.73 |
| Married (lived with partner/ husband/ wife) | 15356 (77.5) | 614 (76.4) | 7306 (77.6) | 7436 (77.5) |  |
| Not married | 4466 (22.5) | 190 (23.6) | 2112 (22.4) | 2164 (22.5) |  |
| Education, n (%) |  |  |  |  | <0.001 |
| Below High School | 11144 (56.2) | 535 (66.5) | 5647 (60.0) | 4962 (51.7) |  |
| College and above | 8678 (43.8) | 269 (33.5) | 3771 (40.0) | 4638 (48.3) |  |
| Hypertension, n (%) | 3904 (19.7) | 208 (25.9) | 2187 (23.2) | 1509 (15.7) | <0.001 |
| Diabetes mellitus, n (%) | 471 (2.4) | 28 (3.5) | 287 (3.0) | 156 (1.6) | <0.001 |
| Heart disease, n (%) | 472 (2.4) | 25 (3.1) | 244 (2.6) | 203 (2.1) | 0.04 |
| Cancer, n (%) | 1087 (5.5) | 37 (4.6) | 520 (5.5) | 530 (5.5) | 0.53 |
| Dyslipidemia, n (%) | 2605 (13.1) | 136 (16.9) | 1400 (14.9) | 1069 (11.1) | <0.001 |
| Healthy diet, n (%) | 13433 (67.8) | 38 (4.7) | 4650 (49.4) | 8745 (91.1) | <0.001 |
| Physically active, n (%) | 14459 (72.9) | 106 (13.2) | 5478 (58.2) | 8875 (92.4) | <0.001 |
| Non-current-smoking, n (%) | 18558 (93.6) | 482 (60.0) | 8577 (91.1) | 9499 (98.9) | <0.001 |
| Moderate alcohol consumption, n (%) | 13371 (67.5) | 76 (9.5) | 4883 (51.8) | 8412 (87.6) | <0.001 |
| Healthy body weight (BMI = 18.5 - <25.0 kg/m<sup>2</sup>), n (%) | 7259 (36.6) | 31 (3.9) | 1556 (16.5) | 5672 (59.1) | <0.001 |
| Brain structural markers |  |  |  |  |  |
| TBV<sup>a</sup>, ml, mean [SD] | 1502.8 ± 72.5 | 1494.3 ± 74.5 | 1499.9 ± 72.0 | 1506.4 ± 72.7 | <0.001 |
| GMV<sup>a</sup>, ml, mean [SD] | 795.6 ± 47.9 | 786.4 ± 49.2 | 792.5 ± 48.2 | 799.4 ± 47.3 | <0.001 |
| WMV<sup>a</sup>, ml, mean [SD] | 707.2 ± 40.7 | 707.8 ± 41.9 | 707.4 ± 40.3 | 707.0 ± 41.1 | 0.73 |
| Hippocampus<sup>a</sup>, ml, mean [SD] | 7.70 ± 0.87 | 7.68 ± 0.88 | 7.71 ± 0.88 | 7.70 ± 0.86 | 0.46 |
| WMHV<sup>a</sup>, ml, median [IQR] | 2.61 (1.42-5.24) | 2.94 (1.56-6.02) | 2.77 (1.50-5.47) | 2.44 (1.32-4.92) | <0.001 |

<sup>a</sup> Head-size normalized brain tissue volumes were used to correct head size.
BMI denotes body mass index; Q denotes quartile; TBV denotes total brain volume; GMV denotes gray matter volume; WMV denotes white matter volume; WMHV denotes white matter hyperintensity volume.

### Number of low-risk lifestyle factors and brain structural markers

Figure 1 and Supplementary Table 2 displays the associations between the number of lifestyle factors and neuroimaging markers. In the cross-sectional analyses from the PRECISE, participants who adopted four or five low-risk lifestyle factors had larger TBV (β=0.12, 95%CI: -0.02, 0.26; *p-trend*=0.048), GMV (β=0.16, 95%CI: 0.01, 0.30; *p-trend*=0.047), but decreased WMHV burden (β=-0.35, 95%CI: -0.50, -0.20; *p-trend*<0.001) and lower odds of lacune (Odds Ratio [OR]=0.48, 95%CI: 0.22, 1.08; *p-trend*=0.03), compared with those with zero or one lifestyle factors. To enable intuitive comparisons, we found that one year of age in the study population was associated with a mean difference of -0.06 (95%CI: -0.07, -0.05) in GMV and 0.06 (95%CI: 0.05, 0.07) in WMHV; thus, the observed association comparing 4-5 to 0-1 low-risk factors were equivalent to approximately 2.7 years of aging delay in GMV and 5.8 years of aging delay in WMHV. No significant association was found between the number of low-risk lifestyle factors and hippocampus volume (β_4-5vs.0-1 lifestyle factors_=-0.03, 95% CI: -0.18, 0.11; *p-trend*=0.59). In the prospective analysis from the UKB, the differences were 0.22 (95% CI: 0.16, 0.28; *p-trend*<0.001) for TBV, 0.26 (95% CI: 0.21, 0.32; *p-trend*<0.001) for GMV, 0.08 (95% CI: 0.01, 0.15; *p-trend*=0.001) for WMV, 0.15 (95% CI: 0.08, 0.22; *p-trend*<0.001) for hippocampus volume, and -0.23 (95% CI: -0.29, -0.17; *p-trend*<0.001) for WMHV burden, for those with four or five low-risk lifestyle factors compared with zero or one low-risk lifestyle factors. Similarly, these association estimates were equivalent to those we found in this study population for approximately 2.0-3.8 years of aging delay in the brain volume.

**Figure 1.**
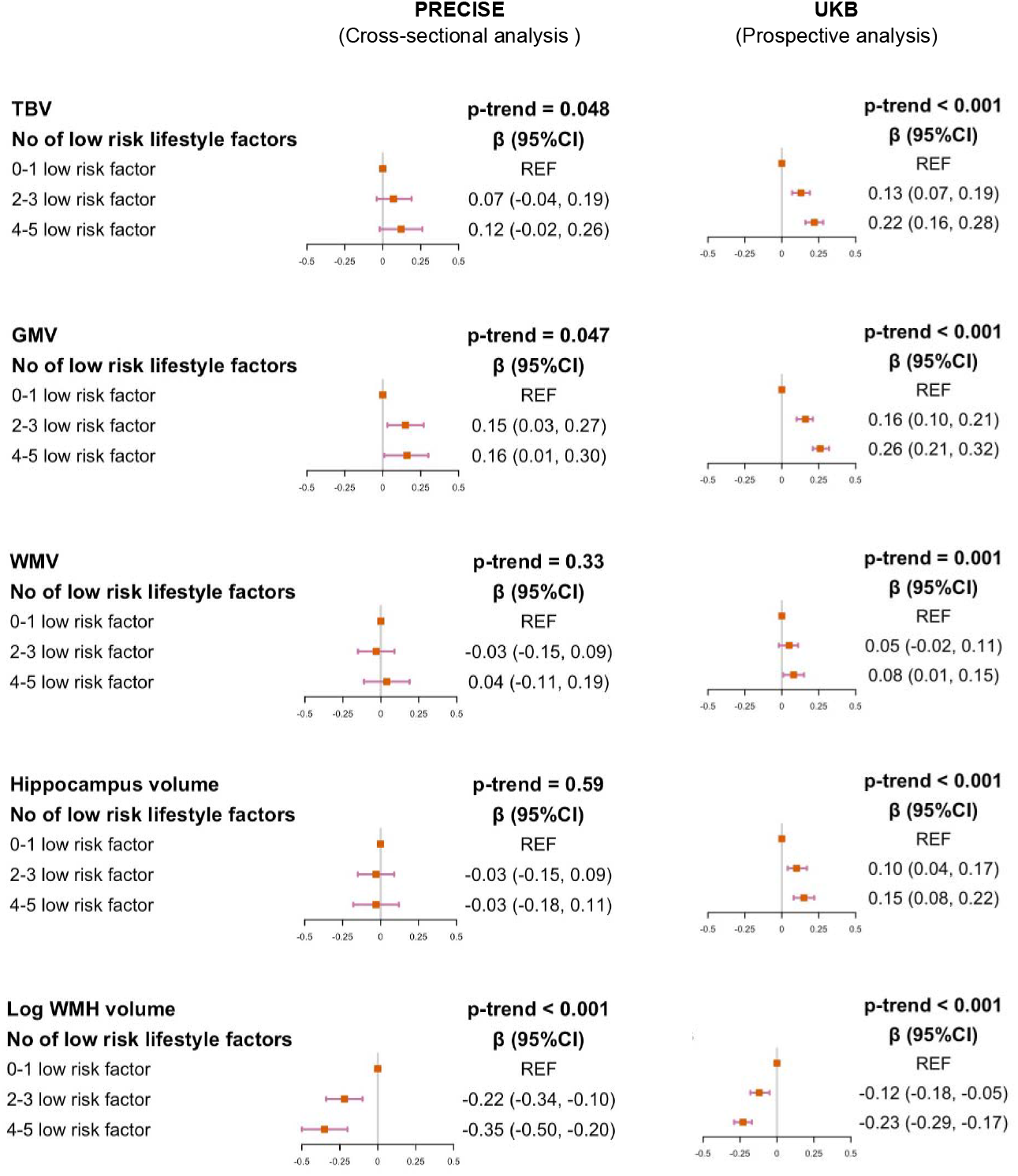
Association between the number of low-risk lifestyle factors and brain MRI markers Multivariable model was adjusted for age at MRI, square of age, gender, ethnicity, marital status, educational levels, and type of residence. For PRECISE, 2413 subjects (302 with 0-1 low-risk lifestyle factors, 1735 with 2-3 low-risk lifestyle factors, 376 with 4-5 low-risk lifestyle factors) were included in the primary analyses; 2408 subjects (300 with 0-1 low-risk lifestyle factors, 1733 with 2-3 low-risk lifestyle factors, 375 with 4-5 low-risk lifestyle factors) included in the analysis of WMH volume. For UKB, 19822 subjects (804 with 0-1 low-risk lifestyle factors, 9418 with 2-3 low-risk lifestyle factors, 9600 with 4-5 low-risk lifestyle factors) were included in the primary analyses. TBV denotes total brain volume; GMV denotes gray matter volume; WMV denotes white matter volume; WMHV denotes white matter hyperintensity volume.

### Individual low-risk lifestyle factors and brain structural markers

In the PRECISE, non-alcohol consumption was associated with larger TBV (β=0.11, 95% CI: 0.01, 0.20; *p-trend*=0.03), GMV (β =0.12, 95% CI: 0.02, 0.22; *p-trend*=0.02), and hippocampus volume (β=0.12, 95% CI: 0.02, 0.22; *p-trend*=0.02); individuals with a healthy body weight had lower hippocampus volume (β=-0.13, 95% CI: -0.20, - 0.05; *p-trend*<0.001) and a decreased WMHV burden (β=-0.33, 95% CI: -0.40, -0.25; *p-trend*<0.001) (Table 3). Meanwhile, physically active, non-current smoking, moderate alcohol consumption and healthy body weight tended to be prospectively associated with a lower degree of a variety of neurodegeneration-related brain structural markers in UKB. Healthy diet was not significantly associated with brain structural markers, except a marginal significant relationship observed with TBV.

**Table 3.** The association between individual low-risk lifestyle factors and brain MRI markers.

| Lifestyle factors | TBV, z-score |  | GMV, z-score |  | WMV, z-score |  | Hippocampus, z-score |  | Log WMHV, z-score |  | Lacune, % |  |
| --- | --- | --- | --- | --- | --- | --- | --- | --- | --- | --- | --- | --- |
|  | B (95% CI) | p | B (95% CI) | p | B (95% CI) | p | B (95% CI) | p | B (95% CI) | p | OR (95% CI) | p |
| **Cross-sectional analysis in PRECISE** |  |  |  |  |  |  |  |  |  |  |  |  |
| Healthy diet | 0.04 (-0.05, 0.13) | 0.41 | 0.02 (-0.07, 0.12) | 0.63 | 0.03 (-0.06, 0.13) | 0.48 | 0.03 (-0.07, 0.12) | 0.57 | 0.02 (-0.08, 0.11) | 0.73 | 0.54 (0.29,1.02) | 0.06 |
| Physically active | 0.02 (-0.07, 0.10) | 0.71 | -0.01 (-0.09, 0.08) | 0.90 | 0.03 (-0.06, 0.12) | 0.54 | 0.03 (-0.06, 0.12) | 0.47 | -0.06 (-0.14, 0.03) | 0.22 | 0.86 (0.51,1.45) | 0.57 |
| Non-current smoking | -0.03 (-0.13, 0.08) | 0.64 | -0.01 (-0.12, 0.10) | 0.88 | -0.03 (-0.14, 0.08) | 0.62 | -0.07 (-0.18, 0.04) | 0.21 | 0.04 (-0.07, 0.15) | 0.47 | 0.62 (0.36,1.07) | 0.08 |
| Non-alcohol consumption | **0.11 (0.01, 0.20)** | **0.03** | **0.12 (0.02, 0.22)** | **0.02** | 0.05 (-0.05, 0.16) | 0.31 | **0.12 (0.02, 0.22)** | **0.02** | -0.05 (-0.16, 0.05) | 0.31 | 1.10 (0.65,1.86) | 0.74 |
| Healthy body weight | 0.04 (-0.03, 0.11) | 0.21 | 0.06 (-0.01, 0.14) | 0.09 | 0.01 (-0.07, 0.08) | 0.83 | **-0.13 (-0.20, -0.05)** | **<0.00<sub>1</sub>** | **-0.33 (-0.40, -0.25)** | **<0.00<sub>1</sub>** | 0.80 (0.53,1.20) | 0.28 |
| **Prospective analysis in UKB** |  |  |  |  |  |  |  |  |  |  |  |  |
| Healthy diet | **-0.03 (-0.05, 0.00)** | **0.05** | -0.02 (-0.04, 0.00) | 0.12 | 0.02 (-0.00, 0.05) | 0.10 | -0.02 (-0.05, 0.01) | 0.12 | -0.01 (-0.04, 0.02) | 0.40 | / | / |
| Physically active | **0.04 (0.01, 0.06)** | **0.01** | **0.03 (0.01, 0.05)** | **0.01** | **0.05 (0.02, 0.07)** | **0.00<sub>3</sub>** | **0.03 (0.00, 0.06)** | **0.04** | -0.02 (-0.04, 0.01) | 0.22 | / | / |
| Non-current smoker | **0.12 (0.07, 0.17)** | **<0.00<sub>1</sub>** | **0.15 (0.11, 0.20)** | **<0.00<sub>1</sub>** | **0.11 (0.05, 0.16)** | **<0.00<sub>1</sub>** | 0.03 (-0.02, 0.09) | 0.249 | **-0.19 (-0.24, -0.14)** | **<0.00<sub>1</sub>** | / | / |
| Moderate alcohol consumption | **0.15 (0.12, 0.17)** | **<0.00<sub>1</sub>** | **0.14 (0.12, 0.17)** | **<0.00<sub>1</sub>** | **0.05 (0.02, 0.08)** | **<0.00<sub>1</sub>** | **0.09 (0.06, 0.12)** | **<0.00<sub>1</sub>** | **-0.06 (-0.09, -0.03)** | **<0.00<sub>1</sub>** | / | / |
| Healthy body weight | **0.06 (0.04, 0.09)** | **<0.00<sub>1</sub>** | **0.10 (0.08, 0.13)** | **<0.00<sub>1</sub>** | -0.01 (-0.04, 0.01) | 0.36 | -0.01 (-0.04, 0.02) | 0.38 | **-0.14 (-0.17, -0.12)** | **<0.00<sub>1</sub>** | / | / |

Multivariable model was adjusted for age at MRI, square of age, gender, Ethnicity, marital status, educational levels, type of residence, and the other four lifestyle variables.
TBV denotes total brain volume; GMV denotes gray matter volume; WMV denotes white matter volume; WMHV denotes white matter hyperintensity volume.

### Subgroup and sensitivity analyses

In PRECISE, we did not observe any significant interactions with age, gender, and education (Supplementary Table 3). The associations were similar across those major subgroups. However, in UKB, we observed stronger associations for GMV and WMHV in females than in males (P for interactions<0.001) (Supplementary Table 4). In addition, the association for WMVH persisted in younger participants (<65 years) but was null in older individuals (P for interaction=0.007).

Multiple sensitivity analyses demonstrated the robustness of our findings. The results of the associations between the number of low-risk lifestyle factors and brain structural markers remained generally unchanged, when we additionally adjusted for the history of major comorbidities, excluded participants with history of major comorbidities, used alternative modifiable healthy lifestyle score by summing up four healthy factors without alcohol factor, used the modifiable healthy lifestyle scores after redefining healthy diet (only in PRECISE) or healthy body weight (only in UKB) (Supplementary Table 5 and Supplementary Table 6). Similar results were found between individual low-risk lifestyle factors and brain structural markers when further adjusted for the history of comorbidities (Supplementary Table 7). When we reconsidered the low-risk of alcohol consumption as non-alcohol consumption in UKB, a significant but attenuated relationship with GMV was observed; however, associations were no longer significant in other brain structural markers (Supplementary Table 8). Furthermore, we found that the number of low-risk lifestyle factors was positively associated with cognitive performance assessed by MoCA score in PRECISE (Supplementary Table 9).

## Discussion

In the cross-sectional study in PRECISE and prospective study in UKB, we observed that adherence to a healthy lifestyle was associated with a panel of major neurodegeneration-related brain structural measures in middle-aged and older adults. Compared with individuals with zero or one low-risk lifestyle factors, those adopted four or five lifestyle factors had larger TBV and GMV and lower WMHV, which was equivalent to approximately 2.0-5.8 years of delay in aging of brain structure. The major contributors were non-alcohol consumption and healthy body weight in PRECISE and physical activity, non-current smoking, moderate alcohol consumption and healthy body weight in UKB.

To our knowledge, the association of overall healthy lifestyle with brain structure has been less explored. Our results are generally consistent with a few recent studies showing that diverse healthy scores (including lifestyle factors, metabolic and health conditions factors) were associated with a variety of brain structure makers^19 20 36^. For example, the Maastricht Study found that middle and older adults who had higher LIBRA (Lifestyle for Brain Health) score (five lifestyle- and seven health-based factors), donating higher dementia risk, were associated with larger WMHV (β_linear_=0.051, *p*=0.002)^19^. The inverse relationship between LIBRA index and GMV has been observed in men, despite a null association observed in the general sample. Similarly, a recent study in UKB study have showed the relation of aggregate vascular risk factors (three lifestyle- and four health-based factors) with lower gray matter volume and higher WMH^36^. Moreover, a study in Spain showed that higher CAIDE (Cardiovascular Risk Factors, Aging, and Incidence of Dementia) Risk Score, including three demographic characteristics, two lifestyle factors, and two health conditions, were associated with white matter hyperintensity load^20^. Compared to those studies, the current study focused particularly on the overall role of modifiable lifestyle factors to better inform targeted public health recommendations regarding primary lifestyle preventions of dementia. Therefore, our study extended previous evidence by elucidating the cross-sectional and prospective associations of five modifiable lifestyle factors with regard to brain structure. Although relatively younger populations included in UKB (mean age 54.83±7.47 years) compared to some previous studies, the consistency of findings across the current two independent studies and the careful control of potential confounding factors suggested that overall lifestyle factors may play a true biological role in maintaining brain structure.

Several individual lifestyle factors showed associations with a panel of brain structural makers, which generally had similar estimates as reported in previous studies. In particular, non-current smoking^9-11^, light to moderate consumption^12 13^/non-alcohol-consumption^14 15^, physical activity^16^ and healthy body weight^17 18^ were related to less brain atrophy (i.e., larger GMV) in previous studies. However, findings investigating the relations of individual risk factors to brain structural markers were not completely consistent between PRECISE and UKB or in most studies, perhaps owing to the differences in study design, sample size, variation of lifestyle patterns across different populations, random error, and reverse causation. For example, we observed an inverse association of healthy body weight with hippocampus volume in PRECISE, whereas a null association in UKB. Nonetheless, results of previous studies have also been mixed with either an inverse association^37^, or a positive association^38^. In addition, we did not observe any protective associations of healthy diet with most brain structure markers in two studies, with the exception of a marginal inverse relationship observed with TBV in UK population, possibly due to the suboptimal dietary assessment methods in both studies. Nevertheless, results from previous studies regarding healthy diet on neuroimaging markers were inconclusive^7 8 39 40^. We observed differences in sex distribution across lifestyle score groups in both studies (72.6% female with four or five low-risk factors group in PRECISE and 41.4% in UKB). This may be explained to a certain extent by sex differences of specific low-risk lifestyle factors between Western and Asian populations, such as non-current smoking and non-alcohol consumption^41 42^. Taken together, further studies are warranted to elucidate the association of healthy lifestyle factors with brain health, either individually or in combination.

Several mechanisms have been proposed to explain the association of overall healthy lifestyle with delaying ageing-related brain atrophy. Possibly, cigarette smoking may act via atherosclerotic processes, which in turn accelerate brain aging^43^. Further, direct toxic effects of smoking may damage the cerebrovascular system, with a concomitant reduction in oxidative imbalances^44^. Greater engagement in physical activity was reported to attenuate the negative association of elevated Aβ burden with cognitive decline and brain atrophy^45^. Taking into account a potential protective role in the upregulation of neurotrophic factors, physical activity may also impact neuronal connectivity and use-dependent plasticity^6^. Moreover, elevated midlife BMI was associated with amyloid deposition in brain, indicating high risk of developing dementia^46^. In addition, given that lifestyle factors are often interrelated, their combination may insert a synergistic influence on brain health^4^.

Major strengths of this study are the large sample size and the availability of individual-level imaging data from two well-established population-based studies in China and UK. In particular, the use of the unprecedented large sample of neuroimaging data in China expanded the existing study scope to the possibly largest ageing society in the world. The availability of a large sample also allows deeper analyses into the specific lifestyle factor accounting for this finding. While inherent difference existed in study design, selected population, criteria of lifestyle, and brain MRI measurement between two studies, we elaborated the robustness of the findings after performing a serious of sensitivity analyses. Nevertheless, some limitations should be noted. First, the cross-sectional nature of PRECISE and may limit the possibility of causal inference between exposures and outcomes. Since we were not able to adjust the baseline brain structures in UKB, the possibility of reverse causation may be inevitable. Second, information of lifestyle factors was self-reported which may lead to potential misclassification. However, non-differential misclassification of a dichotomous exposure may bias the observed associations toward the null. Third, because we lacked the walking data in PRECISE, the daily metabolic equivalent hours of physical activity may lead to underestimation. Forth, given the nature of observational studies, residual confounding may still not be fully eliminated, even though we adjusted extensively for potential risk factors of brain health. Last, although the study population consisted of UK and Chinese populations with nationally representative samples, cautions should be taken when generalizing our findings to other populations.

In summary, our analyses of two independent population-based studies supported a potential beneficial role of an overall healthy lifestyle in maintaining better brain structural health, as manifested by markers of neurodegeneration. Specifically, adherence to a healthier lifestyle was positively associated with total brain volume and gray matter volume, and inversely associated with white matter hyperintensities in middle-aged and older adults. Further large-scale longitudinal studies across different populations are warranted to confirm the study findings and guide public health programs for brain health promotion.

## Supporting information

Supplemental STROBE

## Data Availability

Data of PRECISE are available upon reasonable request. Data are available to researchers on request for purposes of reproducing the results or replicating the procedure by directly contacting the corresponding author. Data from UK Biobank are available on application at www.ukbiobank.ac.uk/register-apply.

## Acknowledgments

We are grateful to all cooperating organizations and their staff in PRECISE and UKB teams whose hard work made this study possible. We thank the interviewees and their families for their voluntary participation in the PRECISE and UKB study.

## Author Contributions

Y.P., C.Y., and J.S. contributed to the conception and design of the study; H.C., G.Z. and A.J performed the statistical analyses; Y.P., X.C., Yi.W., X.M. and Yo.W. administered the PRECISE study. W.Z., J.J. and T.L. processed the imaging data of the PRECISE study. J.S., Y.P., C.Y. and Yo.W. interpreted the data; Y.P., C.Y., and Yo.W. supervised the data analysis and interpretation; J.S. and Y.P. drafted the manuscript; Y.P., C.Y. X.C. and Yo.W. critically reviewed and revised the manuscript; Yo.W. and C.Y. had the primary responsibility for the final content. All authors critically reviewed the manuscript and approved the final draft.

## Data availability statement

For PRECISE study, data are available upon reasonable request. Data are available to researchers on request for purposes of reproducing the results or replicating the procedure by directly contacting the corresponding author. For UKB, data and materials are available via UK Biobank upon application at http://www.ukbiobank.ac.uk/.

## Transparency declaration

The lead author (Y.P.) affirms that the manuscript is an honest, accurate, and transparent account of the study being reported; that no important aspects of the study have been omitted; and that any discrepancies from the study as planned (and, if relevant, registered) have been explained.

## Ethical approval

The study protocol for the PRECISE study was approved by ethics committee at Beijing Tiantan Hospital (IRB approval number: KY2017-010-01) and ethics committee at Lishui Hospital (IRB approval number: 2016-42). The Northwest Multi-Center Research Ethics Committee approved the collection and use of UK Biobank data. Written informed consents were provided from all participants.

