## Supplemental Tables and Figures for "Adherence to a healthy lifestyle and brain structural imaging markers"

**Supplementary Appendix**

### Supplementary Tables and Figures

#### Supplementary Table 1 Definition of low-risk lifestyle factors

| **Lifestyle factors** | **Definition of low-risk lifestyle factor (score = 1)** | |
| --- | --- | --- |
|  | **PRECISE** | **UKB** |
| Diet | **Healthy diet**, assessed by adherence to a diversified diet and defined as in the top 20% of cohort distribution, indicating higher adherence to dietary diversity score (DDS) = [4,5,6] | **Healthy diet**, defined as meeting at least 4 of 7 food groups recommendations:  1. Fruits: ≥ 3 servings/day  2. Vegetables: ≥ 3 servings/day  3. Fish: ≥2 servings/week  4. Processed meats: ≤ 1 serving/week  5. Unprocessed red meats: ≤ 1.5 servings/week  6. Whole grains: ≥ 3servings/day  7. Refined grains: ≤1.5servings/day |
| Physical activity | **Physically active**, defined as the daily vigorous or moderate physical activity (MET-h/d) in the upper quartile of distribution (Q4) | **Physically active**, defined as:  ≥150 minutes moderate activity per week  OR ≥ 75 minutes vigorous activity per week  OR equivalent combination  OR moderate physical activity at least 5 days a week or vigorous activity once a week |
| Smoking status | **Non-current smoking**, defined as never smoker and former smoker | **Non-current smoking**, defined as never smoker and former smoker |
| Alcohol consumption | **Non-alcohol consumption**, defined as never drinker and former drinker | **Moderate alcohol consumption**, defined as >0-14 g/d for women or >0-28 g/d for men |
| Body weight | **Healthy body weight,** defined as BMI in the range of 18.5 - 24.0 kg/m^2^ | **Healthy body weight** was defined as BMI in the range of 20.0 - <25.0 kg/m^2^ |

#### Supplementary Table 2 The association between the number of low-risk lifestyle factors and brain MRI markers

| **Variable** | **TBV, z-score** | | **GMV, z-score** | | **WMV, z-score** | | **Hippocampus volume, z-score** | | **Log WMHV, z-score** | | **Lacune, %** | |
| --- | --- | --- | --- | --- | --- | --- | --- | --- | --- | --- | --- | --- |
|  | **B (95% CI)** | **p** | **B (95% CI)** | **p** | **B (95% CI)** | **p** | **B (95% CI)** | **p** | **B (95% CI)** | **p** | **OR (95% CI)** | **p** |
| **Cross-sectional analysis in PRECISE** | | | | | | | | | | | | |
| **Model 0** |  |  |  |  |  |  |  |  |  |  |  |  |
| 0-1 low-risk factor | REF |  | REF |  | REF |  | REF |  | REF |  | REF |  |
| 2-3 low-risk factor | 0.10 (-0.02, 0.22) | 0.11 | 0.14 (0.02, 0.26) | 0.02 | 0.02 (-0.10, 0.14) | 0.73 | 0.01 (-0.12, 0.13) | 0.91 | -0.10 (-0.22, 0.02) | 0.12 | 0.58 (0.35,0.94) | 0.03 |
| 4-5 low-risk factor | 0.28 (0.13, 0.44) | <0.001 | 0.25 (0.10, 0.40) | 0.001 | 0.20 (0.05, 0.35) | 0.01 | 0.12 (-0.03, 0.27) | 0.12 | -0.28 (-0.43, -0.13) | <0.001 | 0.35 (0.16,0.75) | 0.007 |
| Score, continuous | 0.09 (0.04, 0.13) | <0.001 | 0.07 (0.03, 0.11) | <0.001 | 0.06 (0.02, 0.11) | 0.002 | 0.03 (-0.01, 0.07) | 0.12 | -0.09 (-0.13, -0.05) | <0.001 | 0.73 (0.60,0.89) | 0.002 |
| **Model 1** |  |  |  |  |  |  |  |  |  |  |  |  |
| 0-1 low-risk factor | REF |  | REF |  | REF |  | REF |  | REF |  | REF |  |
| 2-3 low-risk factor | 0.08 (-0.03, 0.19) | 0.16 | 0.17 (0.05, 0.29) | 0.007 | -0.03 (-0.15, 0.09) | 0.65 | -0.02 (-0.14, 0.10) | 0.70 | -0.21 (-0.33, -0.09) | <0.001 | 0.67 (0.39,1.13) | 0.13 |
| 4-5 low-risk factor | 0.14 (-0.002, 0.28) | 0.05 | 0.19 (0.04, 0.33) | 0.02 | 0.04 (-0.11, 0.19) | 0.62 | -0.02 (-0.16, 0.13) | 0.84 | -0.32(-0.47, -0.18) | <0.001 | 0.48 (0.22,1.08) | 0.08 |
| Score, continuous | 0.04 (0.01, 0.08) | 0.02 | 0.05 (0.01, 0.09) | 0.02 | 0.02 (-0.02, 0.06) | 0.33 | -0.01 (-0.05, 0.04) | 0.79 | -0.10 (-0.14, -0.06) | <0.001 | 0.79 (0.64,0.97) | 0.03 |
| **Model 2** | | | | | | | | | | | | |
| 0-1 low-risk factor | REF |  | REF |  | REF |  | REF |  | REF |  | REF |  |
| 2-3 low-risk factor | 0.07 (-0.04, 0.19) | 0.19 | 0.15 (0.03, 0.27) | 0.01 | -0.03 (-0.15, 0.09) | 0.67 | -0.03 (-0.15, 0.09) | 0.62 | -0.22 (-0.34, -0.10) | <0.001 | 0.66 (0.38,1.13) | 0.13 |
| 4-5 low-risk factor | 0.12 (-0.02, 0.26) | 0.09 | 0.16 (0.01, 0.30) | 0.04 | 0.04 (-0.11, 0.19) | 0.63 | -0.03 (-0.18, 0.12) | 0.66 | -0.35 (-0.50, -0.20) | <0.001 | 0.48 (0.22,1.08) | 0.08 |
| Score, continuous | 0.04 (0.00, 0.08) | 0.048 | 0.04 (0.00, 0.08) | 0.047 | 0.02 (-0.02, 0.06) | 0.33 | -0.01 (-0.05, 0.03) | 0.59 | -0.11 (-0.15, -0.07) | <0.001 | 0.79 (0.63,0.98) | 0.03 |
| **Prospective analysis in UKB** | | | | | | | | | | | | |
| **Model 0** |  |  |  |  |  |  |  |  |  |  |  |  |
| 0-1 low-risk factor | REF |  | REF |  | REF |  | REF |  | REF |  | / | / |
| 2-3 low-risk factor | 0.08 (0.01, 0.15) | 0.03 | 0.13 (0.06, 0.20) | <0.001 | -0.01 (-0.08, 0.06) | 0.735 | 0.03 (-0.04, 0.10) | 0.43 | -0.07 (-0.14, 0.00) | 0.05 | / | / |
| 4-5 low-risk factor | 0.17 (0.10, 0.24) | <0.001 | 0.26 (0.20, 0.34) | <0.001 | -0.02 (-0.09, 0.05) | 0.591 | 0.02 (-0.05, 0.09) | 0.60 | -0.18 (-0.25, -0.11) | <0.001 | / | / |
| Score, continuous | 0.05 (0.04, 0.07) | <0.001 | 0.08 (0.07, 0.10) | <0.001 | -0.00 (-0.02, 0.01) | 0.664 | -0.00 (-0.01, 0.01) | 0.79 | -0.06 (-0.07, -0.05) | <0.001 | / | / |
| **Model 1** |  |  |  |  |  |  |  |  |  |  |  |  |
| 0-1 low-risk factor | REF |  | REF |  | REF |  | REF |  | REF |  | / | / |
| 2-3 low-risk factor | 0.11 (0.07, 0.18) | <0.001 | 0.15 (0.10, 0.21) | <0.001 | 0.04 (-0.03, 0.11) | 0.23 | 0.11 (0.04, 0.17) | 0.001 | -0.12 (-0.18, -0.06) | <0.001 | / | / |
| 4-5 low-risk factor | 0.21 (0.15, 0.27) | <0.001 | 0.26 (0.20, 0.31) | <0.001 | 0.07 (0.00, 0.14) | 0.04 | 0.17 (0.10, 0.23) | <0.001 | -0.24 (-0.30, -0.17) | <0.001 | / | / |
| Score, continuous | 0.06 (0.04, 0.07) | <0.001 | 0.07 (0.06, 0.08) | <0.001 | 0.02 (0.01, 0.03) | 0.004 | 0.04 (0.03, 0.05) | <0.001 | -0.07 (-0.08, -0.06) | <0.001 | / | / |
| **Model 2** |  |  |  |  |  |  |  |  |  |  |  |  |
| 0-1 low-risk factor | REF |  | REF |  | REF |  | REF |  | REF |  | / | / |
| 2-3 low-risk factor | 0.13 (0.07, 0.19) | <0.001 | 0.16 (0.10, 0.21) | <0.001 | 0.05 (-0.02, 0.11) | 0.19 | 0.10 (0.04, 0.17) | 0.002 | -0.12 (-0.18, -0.05) | <0.001 | / | / |
| 4-5 low-risk factor | 0.22 (0.16, 0.28) | <0.001 | 0.26 (0.21, 0.32) | <0.001 | 0.08 (0.01, 0.15) | 0.027 | 0.15 (0.08, 0.22) | <0.001 | -0.23 (-0.29, -0.17) | <0.001 | / | / |
| Score, continuous | 0.06 (0.05, 0.07) | <0.001 | 0.07 (0.06, 0.08) | <0.001 | 0.02 (0.01, 0.03) | 0.001 | 0.03 (0.02, 0.04) | <0.001 | -0.07 (-0.08, -0.06) | <0.001 | / | / |

Model 0 was crude model. Model 1 was adjusted for age at MRI, square of age, gender. Model 2 was additionally adjusted for ethnicity, marital status, educational levels, and type of residence.

TBV denotes total brain volume; GMV denotes gray matter volume; WMV denotes white matter volume; WMHV denotes white matter hyperintensity volume.

#### Supplementary Table 3 Subgroup analysis for the cross-sectional association of the number of low-risk lifestyle factors with brain MRI markers in PRECISE, stratified by age, gender, and education

| **Subgroups** | **No. (%)** | **TBV, z-score** | | **GMV, z-score** | | **WMV, z-score** | | **Hippocampus volume, z-score** | | **Log WMHV, z-score** | | **Lacune, %** | |
| --- | --- | --- | --- | --- | --- | --- | --- | --- | --- | --- | --- | --- | --- |
|  |  | **B (95% CI)** | **p** | **B (95% CI)** | **p** | **B (95% CI)** | **p** | **B (95% CI)** | **p** | **B (95% CI)** | **p** | **OR (95% CI)** | **p** |
| **Age** |  |  |  |  |  |  |  |  |  |  |  |  |  |
| <65 y | 1682 (69.7) | 0.05 (0.00, 0.09) | 0.048 | 0.05 (-0.00, 0.09) | 0.05 | 0.03 (-0.02, 0.07) | 0.30 | 0.00 (-0.04, 0.05) | 0.86 | -0.10 (-0.15, -0.05) | <0.001 | 0.79 (0.58, 1.08) | 0.14 |
| >=65 y | 731 (30.3) | 0.02 (-0.05, 0.10) | 0.55 | 0.04 (-0.04, 0.12) | 0.34 | -0.00 (-0.08, 0.08) | 0.96 | -0.04 (-0.12, 0.04) | 0.31 | -0.11 (-0.18, -0.04) | 0.001 | 0.81 (0.60, 1.10) | 0.17 |
| P for interaction |  |  | 0.74 |  | 0.75 |  | 0.87 |  | 0.51 |  | 0.56 |  | 0.42 |
| **Gender** |  |  |  |  |  |  |  |  |  |  |  |  |  |
| Male | 1113 (46.1) | 0.05 (-0.01, 0.10) | 0.09 | 0.06 (0.00, 0.11) | 0.049 | 0.02 (-0.04, 0.07) | 0.60 | -0.02 (-0.08, 0.04) | 0.48 | -0.09 (-0.14, -0.04) | <0.001 | 0.77 (0.59, 0.99) | 0.04 |
| Female | 1300 (53.9) | 0.03 (-0.03, 0.09) | 0.29 | 0.03 (-0.03, 0.08) | 0.41 | 0.02 (-0.04, 0.08) | 0.45 | 0.02 (-0.04, 0.08) | 0.60 | -0.13 (-0.20, -0.07) | <0.001 | 0.87 (0.57, 1.33) | 0.52 |
| P for interaction |  |  | 0.74 |  | 0.42 |  | 0.80 |  | 0.48 |  | 0.23 |  | 0.64 |
| **Education** |  |  |  |  |  |  |  |  |  |  |  |  |  |
| Illiteracy | 387 (16.0) | 0.07 (-0.03, 0.17) | 0.16 | 0.08 (-0.03, 0.19) | 0.15 | 0.03 (-0.07, 0.14) | 0.50 | 0.03 (-0.08, 0.14) | 0.58 | -0.12 (-0.23, -0.00) | 0.049 | 0.90 (0.52, 1.54) | 0.69 |
| Literacy | 2026 (84.0) | 0.04 (-0.01, 0.08) | 0.10 | 0.04 (-0.01, 0.08) | 0.10 | 0.02 (-0.03, 0.06) | 0.41 | -0.02 (-0.06,0.03) | 0.45 | -0.10 (-0.14, -0.06) | <0.001 | 0.78 (0.61, 0.99) | 0.04 |
| P for interaction |  |  | 0.65 |  | 0.56 |  | 0.90 |  | 0.46 |  | 0.67 |  | 0.49 |

Adjusted covariates included age at MRI, square of age, gender, ethnicity, marital status, educational levels, type of residence.

TBV denotes total brain volume; GMV denotes gray matter volume; WMV denotes white matter volume; WMHV denotes white matter hyperintensity volume.

#### Supplementary Table 4 Subgroup analysis for the prospective association of the number of low-risk lifestyle factors with brain MRI markers in UKB, stratified by age, gender, and education

| **Subgroups** | **No. (%)** | **TBV, z-score** | | **GMV, z-score** | | **WMV, z-score** | | **Hippocampus volume, z-score** | | **Log WMHV, z-score** | |
| --- | --- | --- | --- | --- | --- | --- | --- | --- | --- | --- | --- |
|  |  | **B (95% CI)** | **p** | **B (95% CI)** | **p** | **B (95% CI)** | **p** | **B (95% CI)** | **p** | **B (95% CI)** | **p** |
| **Age** |  |  |  |  |  |  |  |  |  |  |  |
| <65 y | 17878 | 0.06 (0.05, 0.07) | <0.001 | 0.07 (0.06, 0.08) | <0.001 | 0.02 (0.01, 0.04) | <0.001 | 0.03 (0.02, 0.05) | <0.001 | -0.08 (-0.09, -0.06) | <0.001 |
| >=65 y | 1944 | 0.04 (0.00, 0.08) | 0.03 | 0.07 (0.03, 0.10) | <0.001 | -0.01 (-0.05, 0.04) | 0.78 | 0.03 (-0.01, 0.07) | 0.18 | 0.01 (-0.03, 0.05) | 0.52 |
| P for interaction |  |  | 0.06 |  | 0.92 |  | **0.005** |  | 0.80 |  | **0.007** |
| **Gender** |  |  |  |  |  |  |  |  |  |  |  |
| Male | 10426 | 0.04 (0.02, 0.05) | <0.001 | 0.04 (0.03, 0.06) | <0.001 | 0.01 (-0.01, 0.03) | 0.16 | 0.03 (0.02, 0.05) | <0.001 | -0.05 (-0.07, -0.03) | <0.001 |
| Female | 9396 | 0.08 (0.07, 0.10) | <0.001 | 0.10 (0.08, 0.11) | <0.001 | 0.03 (0.01, 0.05) | 0.001 | 0.03 (0.01, 0.05) | 0.004 | -0.09 (-0.10, -0.07) | <0.001 |
| P for interaction |  |  | **<0.001** |  | **<0.001** |  | 0.12 |  | 0.46 |  | **<0.001** |
| **Education** |  |  |  |  |  |  |  |  |  |  |  |
| High School and Below | 11144 | 0.05 (0.04, 0.07) | <0.001 | 0.07 (0.05, 0.08) | <0.001 | 0.01 (-0.00, 0.03) | 0.16 | 0.04 (0.02, 0.05) | <0.001 | -0.07 (-0.09, -0.06) | <0.001 |
| College and Above | 8678 | 0.07 (0.05, 0.09) | <0.001 | 0.08 (0.06, 0.09) | <0.001 | 0.03 (0.01, 0.05) | 0.001 | 0.03 (0.01, 0.05) | 0.01 | -0.06 (-0.08, -0.04) | <0.001 |
| P for interaction |  |  | 0.31 |  | 0.60 |  | **0.04** |  | 0.97 |  | 0.12 |

Adjusted covariates included age at MRI, square of age, gender, ethnicity, marital status, educational levels, type of residence.

TBV denotes total brain volume; GMV denotes gray matter volume; WMV denotes white matter volume; WMHV denotes white matter hyperintensity volume.

#### Supplementary Table 5 Sensitivity analysis for the association of the number of low-risk lifestyle factors with brain MRI markers in PRECISE

| **Variable** | **TBV, z-score** | | **GMV, z-score** | | **WMV, z-score** | | **Hippocampus volume, z-score** | | **Log WMHV, z-score** | | **Lacune, %** | |
| --- | --- | --- | --- | --- | --- | --- | --- | --- | --- | --- | --- | --- |
|  | **B (95% CI)** | **p** | **B (95% CI)** | **p** | **B (95% CI)** | **p** | **B (95% CI)** | **p** | **B (95% CI)** | **p** | **OR (95% CI)** | **p** |
| **Sensitivity analysis 1*****: Multivariable analysis after additional adjustment for history of major comorbidities, including hypertension, heart disease, diabetes mellitus, tumors, or dyslipidemia** | | | | | | | | | | | | |
| 0-1 low-risk factor | REF |  | REF |  | REF |  | REF |  | REF |  | REF |  |
| 2-3 low-risk factor | 0.07 (-0.04, 0.19) | 0.19 | 0.15 (0.03, 0.27) | 0.01 | -0.02 (-0.14, 0.10) | 0.71 | -0.02 (-0.14, 0.10) | 0.73 | -0.20 (-0.31, -0.08) | 0.001 | 0.71 (0.41,1.21) | 0.21 |
| 4-5 low-risk factor | 0.10 (-0.04, 0.24) | 0.16 | 0.13 (-0.02, 0.28) | 0.09 | 0.03 (-0.12, 0.18) | 0.68 | -0.02 (-0.17, 0.13) | 0.81 | -0.29 (-0.44, -0.14) | <0.001 | 0.57 (0.25,1.28) | 0.17 |
| Score, continuous | 0.03 (-0.01, 0.07) | 0.10 | 0.03 (-0.01, 0.07) | 0.12 | 0.02 (-0.02, 0.06) | 0.39 | -0.01 (-0.05, 0.04) | 0.79 | -0.09 (-0.13, -0.05) | <0.001 | 0.82 (0.66,1.01) | 0.07 |
| **Sensitivity analysis 2**†**: Excluding participants with history of major comorbidities (N = 785)** | | | | | | | | | | | | |
| 0-1 low-risk factor | REF |  | REF |  | REF |  | REF |  | REF |  | REF |  |
| 2-3 low-risk factor | 0.15 (-0.04, 0.35) | 0.13 | 0.19 (-0.02, 0.40) | 0.07 | 0.06 (-0.15, 0.26) | 0.59 | 0.10 (-0.11, 0.31) | 0.35 | -0.31 (-0.53, -0.09) | 0.01 | 1.28 (0.34,4.74) | 0.72 |
| 4-5 low-risk factor | 0.06 (-0.18, 0.30) | 0.60 | 0.07 (-0.18, 0.32) | 0.59 | 0.03 (-0.21, 0.28) | 0.79 | 0.04 (-0.21, 0.29) | 0.74 | -0.30 (-0.56, -0.03) | 0.03 | 0.92 (0.16,5.20) | 0.93 |
| Score, continuous | 0.04 (-0.03, 0.11) | 0.25 | 0.03 (-0.04, 0.10) | 0.35 | 0.03 (-0.04, 0.10) | 0.42 | 0.04 (-0.03, 0.11) | 0.31 | -0.07 (-0.14, 0.01) | 0.07 | 0.77 (0.49,1.21) | 0.25 |
| **Sensitivity analysis 3**†**: Modified healthy lifestyle score that was based on the other 4 healthy factors without alcohol** | | | | | | | | | | | | |
| 0-1 low-risk factor | REF |  | REF |  | REF |  | REF |  | REF |  | REF |  |
| 2-3 low-risk factor | 0.05 (-0.02, 0.12) | 0.17 | 0.04 (-0.03, 0.12) | 0.26 | 0.03 (-0.04, 0.11) | 0.38 | -0.08 (-0.15, 0.00) | 0.05 | -0.21 (-0.28, -0.13) | <0.001 | 0.54 (0.36,0.82) | 0.004 |
| 4-5 low-risk factor | 0.07 (-0.19, 0.34) | 0.60 | 0.05 (-0.23, 0.34) | 0.72 | 0.06 (-0.22, 0.34) | 0.68 | -0.06 (-0.35, 0.22) | 0.66 | -0.19 (-0.47, 0.09) | 0.19 | 0.88 (0.20,3.85) | 0.86 |
| Score, continuous | 0.03 (-0.01, 0.07) | 0.17 | 0.03 (-0.02, 0.08) | 0.18 | 0.02 (-0.03, 0.06) | 0.47 | -0.04 (-0.08, 0.01) | 0.11 | -0.12 (-0.17, -0.08) | <0.001 | 0.73 (0.57,0.94) | 0.01 |
| **Sensitivity analysis 4**†**: Modified healthy lifestyle score after redefining the healthy diet as those who ate vegetables and fruits every day and red meat 1 to 6 days a week** | | | | | | | | | | | | |
| 0-1 low-risk factor | REF |  | REF |  | REF |  | REF |  | REF |  | REF |  |
| 2-3 low-risk factor | 0.07 (-0.05, 0.18) | 0.25 | 0.08 (-0.04, 0.19) | 0.22 | 0.03 (-0.09, 0.15) | 0.62 | -0.01 (-0.13, 0.11) | 0.89 | -0.22 (-0.34, -0.10) | <0.001 | 0.75 (0.43,1.30) | 0.31 |
| 4-5 low-risk factor | -0.03 (-0.17, 0.11) | 0.66 | 0.01 (-0.14, 0.16) | 0.89 | -0.06 (-0.21, 0.10) | 0.47 | -0.11 (-0.26, 0.04) | 0.15 | -0.40 (-0.55, -0.25) | <0.001 | 0.78 (0.36,1.67) | 0.52 |
| Score, continuous | 0.02 (-0.02, 0.05) | 0.42 | 0.02 (-0.02, 0.07) | 0.23 | 0.001 (-0.04, 0.04) | 0.94 | -0.01 (-0.05, 0.03) | 0.61 | -0.12 (-0.16, -0.08) | <0.001 | 0.87 (0.70,1.09) | 0.23 |

* Model was adjusted for age at MRI, square of age, gender, ethnicity, marital status, educational levels, type of residence, and history of major comorbidities, including hypertension, heart disease, diabetes mellitus, tumors, or dyslipidemia.

† Model was adjusted for age at MRI, square of age, gender, ethnicity, marital status, educational level, type of residence.

TBV denotes total brain volume; GMV denotes gray matter volume; WMV denotes white matter volume; WMHV denotes white matter hyperintensity volume.

#### Supplementary Table 6 Sensitivity analysis for the association of the number of low-risk lifestyle factors with brain MRI markers in UKB

| **Variable** | **TBV, z-score** | | **GMV, z-score** | | **WMV, z-score** | | **Hippocampus volume, z-score** | | **Log WMHV, z-score** | |
| --- | --- | --- | --- | --- | --- | --- | --- | --- | --- | --- |
|  | **B (95% CI)** | **p** | **B (95% CI)** | **p** | **B (95% CI)** | **p** | **B (95% CI)** | **p** | **B (95% CI)** | **p** |
| **Sensitivity analysis 1*****: Multivariable analysis after additional adjustment for history of major comorbidities, including hypertension, heart disease, diabetes mellitus, cancer, or dyslipidemia** | | | | | | | | | | |
| 0-1 low-risk factor | REF |  | REF |  | REF |  | REF |  | REF |  |
| 2-3 low-risk factor | 0.13 (0.07,0.19) | <0.001 | 0.15 (0.10, 0.21) | <0.001 | 0.05 (-0.02, 0.11) | 0.19 | 0.10 (0.04, 0.17) | 0.003 | -0.11 (-0.17, -0.05) | 0.001 |
| 4-5 low-risk factor | 0.21 (0.15,0.27) | <0.001 | 0.25 (0.19, 0.30) | <0.001 | 0.08 (0.01, 0.15) | 0.03 | 0.14 (0.08, 0.21) | <0.001 | -0.20 (-0.27, -0.14) | <0.001 |
| Score, continuous | 0.05 (0.04,0.07) | <0.001 | 0.06 (0.05, 0.07) | <0.001 | 0.02 (0.01, 0.03) | 0.001 | 0.03 (0.02, 0.04) | <0.001 | -0.06 (-0.07, -0.04) | <0.001 |
| **Sensitivity analysis 2**†**: Excluding participants with history of major comorbidities (N = 13609)** | | | | | | | | | | |
| 0-1 low-risk factor | REF |  | REF |  | REF |  | REF |  | REF |  |
| 2-3 low-risk factor | 0.09 (0.02,0.17) | 0.016 | 0.14 (0.07, 0.20) | <0.001 | 0.00 (-0.08, 0.09) | 0.99 | 0.09 (0.01, 0.17) | 0.037 | -0.10 (-0.17, -0.02) | 0.015 |
| 4-5 low-risk factor | 0.17 (0.10,0.25) | <0.001 | 0.23 (0.17, 0.30) | <0.001 | 0.03 (-0.05, 0.12) | 0.44 | 0.13 (0.05, 0.22) | 0.002 | -0.22 (-0.29, -0.14) | <0.001 |
| Score, continuous | 0.05 (0.04,0.07) | <0.001 | 0.06 (0.05, 0.07) | <0.001 | 0.02 (0.00, 0.03) | 0.01 | 0.03 (0.01, 0.04) | <0.001 | -0.06 (-0.08, -0.05) | <0.001 |
| **Sensitivity analysis 3**†**: Modified healthy lifestyle score that was based on the other 4 healthy factors without alcohol** | | | | | | | | | | |
| 0-1 low-risk factor | REF |  | REF |  | REF |  | REF |  | REF |  |
| 2-3 low-risk factor | 0.05 (0.01, 0.09) | 0.02 | 0.07 (0.04, 0.11) | <0.001 | -0.01 (-0.05, 0.04) | 0.83 | 0.07 (0.02, 0.11) | 0.003 | -0.11 (-0.15, -0.07) | <0.001 |
| 4-5 low-risk factor | 0.11 (0.06, 0.15) | <0.001 | 0.16 (0.12, 0.20) | <0.001 | 0.00 (-0.05, 0.06) | 0.91 | 0.09 (0.04, 0.14) | 0.001 | -0.21 (-0.26, -0.16) | <0.001 |
| Score, continuous | 0.04 (0.02, 0.05) | <0.001 | 0.05 (0.04, 0.06) | <0.001 | 0.00 (-0.01, 0.02) | 0.68 | 0.03 (0.01, 0.04) | <0.001 | -0.07 (-0.08, -0.06) | <0.001 |
| **Sensitivity analysis 4**†**: Modified healthy lifestyle score after redefining the healthy body weight as BMI in range of 18.5-24.9 kg/m^2^** | | | | | | | | | | |
| 0-1 low-risk factor | REF |  | REF |  | REF |  | REF |  | REF |  |
| 2-3 low-risk factor | 0.14 (0.08,0.20) | <0.001 | 0.16 (0.11,0.22) | <0.001 | 0.05 (-0.02,0.12) | 0.143 | 0.11 (0.04,0.18) | 0.001 | -0.12 (-0.18, -0.06) | <0.001 |
| 4-5 low-risk factor | 0.22 (0.16,0.28) | <0.001 | 0.27 (0.21,0.32) | <0.001 | 0.08 (0.01,0.15) | 0.024 | 0.15 (0.08,0.21) | <0.001 | -0.24 (-0.30, -0.17) | <0.001 |
| Score, continuous | 0.06 (0.05,0.07) | <0.001 | 0.07 (0.06,0.08) | <0.001 | 0.02 (0.01,0.03) | 0.003 | 0.03 (0.02,0.04) | <0.001 | -0.07 (-0.08, -0.06) | <0.001 |

* Model was adjusted for age at MRI, square of age, gender, ethnicity, marital status, educational levels, type of residence, and history of major comorbidities, including hypertension, heart disease, diabetes mellitus, cancer, or dyslipidemia

† Model was adjusted for age at MRI, square of age, gender, ethnicity, marital status, educational level, type of residence.

BMI denotes body mass index; TBV denotes total brain volume; GMV denotes gray matter volume; WMV denotes white matter volume; WMHV denotes white matter hyperintensity volume.

#### Supplementary Table 7 Sensitivity analysis for the association of individual low-risk lifestyle factors with brain markers, additionally adjustment for history of major comorbidities in PRECISE and UKB

| **Lifestyle factors** | **TBV, z-score** | | **GMV, z-score** | | **WMV, z-score** | | **Hippocampus volume, z-score** | | **Log WMH volume, z-score** | | **Lacune, %** | |
| --- | --- | --- | --- | --- | --- | --- | --- | --- | --- | --- | --- | --- |
|  | **B (95% CI)** | **p** | **B (95% CI)** | **p** | **B (95% CI)** | **p** | **B (95% CI)** | **p** | **B (95% CI)** | **p** | **OR (95% CI)** | **p** |
| **Cross-sectional analysis in PRECISE*** | | | | | | | | | | | | |
| Healthy diet | 0.04 (-0.05, 0.13) | 0.39 | 0.02 (-0.07, 0.12) | 0.61 | 0.03 (-0.06, 0.13) | 0.47 | 0.03 (-0.06, 0.13) | 0.52 | 0.02 (-0.08, 0.11) | 0.74 | 0.53 (0.28, 1.01) | 0.05 |
| Physically active | 0.02 (-0.06, 0.10) | 0.64 | -0.002 (-0.09, 0.09) | 0.97 | 0.03 (-0.06, 0.12) | 0.50 | 0.04 (-0.04, 0.13) | 0.33 | -0.06 (-0.14, 0.03) | 0.21 | 0.86 (0.51, 1.45) | 0.56 |
| Non-current smoker | -0.01 (-0.12, 0.09) | 0.80 | 0.002 (-0.11, 0.11) | 0.98 | -0.02 (-0.13, 0.09) | 0.71 | -0.06 (-0.18, 0.05) | 0.26 | 0.04 (-0.07, 0.15) | 0.50 | 0.63 (0.36, 1.08) | 0.09 |
| Non-current drinker | 0.10 (0.00, 0.19) | 0.049 | 0.11 (0.01, 0.21) | 0.04 | 0.05 (-0.06, 0.15) | 0.38 | 0.11 (0.01, 0.22) | 0.03 | -0.04 (-0.14, 0.06) | 0.44 | 1.16 (0.68, 1.98) | 0.59 |
| Healthy body weight | 0.02 (-0.05,0.09) | 0.60 | 0.03 (-0.05, 0.10) | 0.44 | 0.001 (-0.07, 0.08) | 0.97 | -0.12 (-0.20, -0.05) | 0.001 | -0.28 (-0.36, -0.21) | <0.001 | 0.87 (0.58, 1.32) | 0.51 |
| **Prospective analysis in UKB** † | | | | | | | | | | | | |
| Healthy diet | -0.03 (-0.05, -0.00) | 0.05 | -0.02 (-0.04, 0.00) | 0.11 | -0.02 (-0.05, 0.01) | 0.12 | 0.02 (-0.00, 0.05) | 0.10 | -0.01 (-0.04, 0.02) | 0.41 | / | / |
| Physically active | 0.03 (0.01, 0.06) | 0.02 | 0.02 (-0.00, 0.05) | 0.07 | 0.03 (0.00, 0.06) | 0.04 | 0.04 (0.01, 0.07) | 0.01 | -0.01 (-0.03, 0.02) | 0.63 | / | / |
| Non-current smoker | 0.12 (0.07, 0.17) | <0.001 | 0.16 (0.12, 0.20) | <0.001 | 0.03 (-0.02, 0.09) | 0.26 | 0.11 (0.05, 0.16) | <0.001 | -0.19 (-0.24, -0.14) | <0.001 | / | / |
| Moderate alcohol consumption | 0.15 (0.12, 0.17) | <0.001 | 0.14 (0.12, 0.17) | <0.001 | 0.09 (0.06, 0.12) | <0.001 | 0.05 (0.02, 0.08) | <0.001 | -0.06 (-0.08, -0.03) | <0.001 | / | / |
| Healthy body weight | 0.05 (0.02, 0.07) | <0.001 | 0.08 (0.06, 0.11) | <0.001 | -0.01 (-0.04, 0.02) | 0.38 | -0.02 (-0.05, 0.00) | 0.11 | -0.11 (-0.14, -0.08) | <0.001 | / | / |

* Models were adjusted for age at MRI, square of age, gender, ethnicity, marital status, educational levels, type of residence, the other four lifestyle variables, and history of major comorbidities (hypertension, heart disease, diabetes mellitus, tumors, or dyslipidemia).

† Models were adjusted for age at MRI, square of age, gender, ethnicity, marital status, educational levels, type of residence, the other four lifestyle variables, and history of major comorbidities (hypertension, heart disease, diabetes mellitus, cancer, or dyslipidemia).

TBV denotes total brain volume; GMV denotes gray matter volume; WMV denotes white matter volume; WMHV denotes white matter hyperintensity volume.

#### Supplementary Table 8 Sensitivity analysis for the association of individual low-risk lifestyle factors with brain markers in UKB, after redefining the healthy level of alcohol consumption factor as non-current drinker

| **Lifestyle factors** | **TBV, z-score** | | **GMV, z-score** | | **WMV, z-score** | | **Hippocampus volume, z-score** | | **Log WMH volume, z-score** | |
| --- | --- | --- | --- | --- | --- | --- | --- | --- | --- | --- |
|  | **B (95% CI)** | **p** | **B (95% CI)** | **p** | **B (95% CI)** | **p** | **B (95% CI)** | **p** | **B (95% CI)** | **p** |
| Healthy diet | -0.02 (-0.04, 0.01) | 0.15 | -0.01 (-0.03, 0.01) | 0.32 | -0.02 (-0.05, 0.01) | 0.20 | 0.03 (-0.00, 0.05) | 0.07 | -0.01 (-0.04, 0.01) | 0.29 |
| Physically active | 0.04 (0.01, 0.06) | 0.01 | 0.03 (0.01, 0.05) | 0.02 | 0.03 (-0.00, 0.06) | 0.05 | 0.04 (0.01, 0.07) | 0.004 | -0.02 (-0.04, 0.01) | 0.28 |
| Non-current smoker | 0.14 (0.09, 0.19) | <0.001 | 0.17 (0.13, 0.22) | <0.001 | 0.05 (-0.01, 0.10) | 0.10 | 0.11 (0.06, 0.17) | <0.001 | -0.20 (-0.25, -0.15) | <0.001 |
| Non-current drinker | 0.03 (-0.01, 0.06) | 0.11 | 0.04 (0.01, 0.07) | 0.006 | -0.00 (-0.04, 0.03) | 0.95 | -0.02 (-0.05, 0.01) | 0.27 | 0.03 (-0.00, 0.06) | 0.09 |
| Healthy body weight | 0.06 (0.04, 0.09) | <0.001 | 0.11 (0.08, 0.13) | <0.001 | -0.01 (-0.04, 0.02) | 0.43 | -0.01 (-0.04, 0.02) | 0.38 | -0.14 (-0.17, -0.12) | <0.001 |

Multivariable model was adjusted for age at MRI, square of age, gender, ethnicity, marital status, educational levels, and type of residence.

TBV denotes total brain volume; GMV denotes gray matter volume; WMV denotes white matter volume; WMHV denotes white matter hyperintensity volume.

#### Supplementary Table 9 The association of the number of low-risk lifestyle factors and components with MoCA score in PRECISE

| **Variable** | **Model 0** | | **Model 1** | | **Model 2** | | **Model 3** | |
| --- | --- | --- | --- | --- | --- | --- | --- | --- |
|  | **B (95% CI)** | **p** | **B (95% CI)** | **p** | **B (95% CI)** | **p** | **B (95% CI)** | **p** |
| **Healthy lifestyle score** |  |  |  |  |  |  |  |  |
| 0-1 low-risk factor | REF |  | REF |  | REF |  | REF |  |
| 2-3 low-risk factor | -0.15 (-0.62, 0.33) | 0.54 | -0.07 (-0.58, 0.43) | 0.77 | -0.11 (-0.61, 0.39) | 0.66 | -0.15 (-0.65, 0.35) | 0.56 |
| 4-5 low-risk factor | 0.35 (-0.24, 0.94) | 0.24 | 0.37 (-0.25, 1,00) | 0.24 | 0.32 (-0.31, 0.94) | 0.32 | 0.25 (-0.37, 0.88) | 0.43 |
| Score, continuous | 0.17 (0.01, 0.33) | 0.04 | 0.19 (0.02, 0.36) | 0.03 | 0.18 (0.01, 0.35) | 0.03 | 0.17 (-0.00, 0.34) | 0.05 |

Model 1 was adjusted for age at MRI, square of age, gender. Model 2 was additionally adjusted for ethnicity, marital status, educational levels, type of residence. Model 3 was additionally adjusted for history of major comorbidities, including hypertension, heart disease, diabetes mellitus, tumors, or dyslipidemia.

TBV denotes total brain volume; GMV denotes gray matter volume; WMV denotes white matter volume; WMHV denotes white matter hyperintensity volume.

#### Supplementary Figure 1 Participant selection flowchart

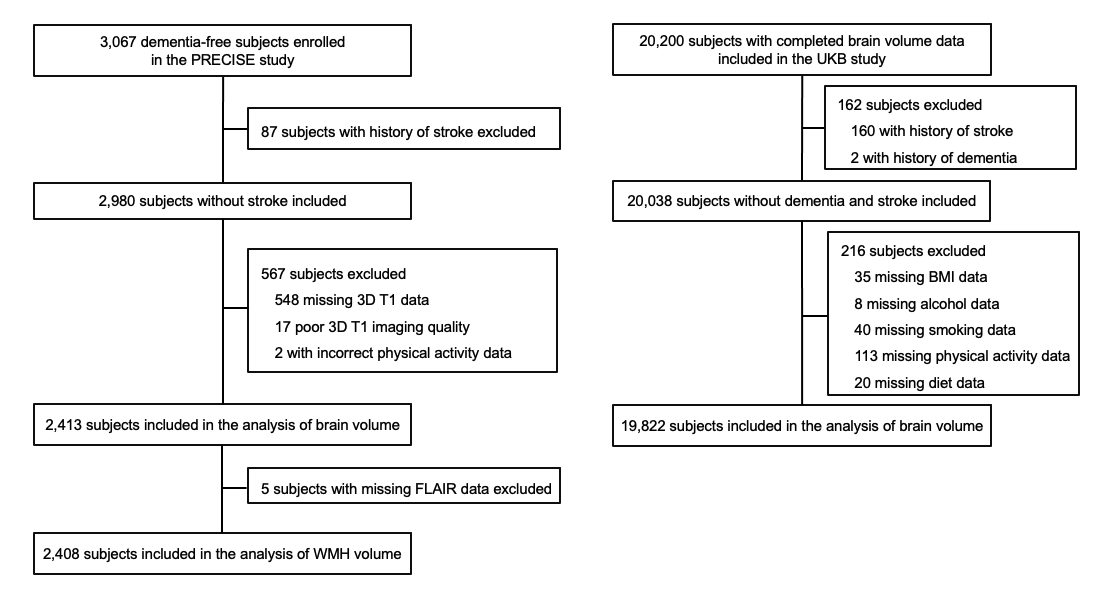
